# Functionalizing face masks with natural clays: preliminary results

**DOI:** 10.1101/2023.07.26.23293200

**Authors:** D. Hernández, L. A. Rodríguez-de-Torner, E. Altshuler, A. Rivera

## Abstract

The materials used in the fabrication of standard, three-layered surgical masks are functionalized by the incorporation of Cuban natural clay with potential microbicidal properties. Different treatments involving aqueous solutions of bentonite (Bent) clay and small amounts of a cationic surfactant are studied. Optical microscopy indicates that the clay particles are effectively adhered to the fibers forming the mask layers. The capacity of the materials to block ballistic droplets showed to be very high for the external and medium layers of a surgical mask both before and after functionalization. However, functionalization demonstrated to increase the blocking capacity of the material in the inner layer, i.e., that closer to the user’s face.

## INTRODUCTION

From the early beginnings of the COVID pandemic, face masks have had a key role in slowing down the transmission of SARS-CoV-2. Many studies assessed masks’ efficacy [1-10], which has compelled health authorities to recommend the use of face masks in certain settings. This includes home-made cloth masks and surgical masks, which are very popular in countries like Cuba.

Evidence suggests that SARS-CoV-2 is basically transmitted through exposure to respiratory droplets carrying the virus [11]. This transmission route is usually divided into two possibilities. The first –*ballistic transmission*– occurs by the emission of large droplets of fluid as infected individuals sneeze, cough, sing, or talk. Those droplets, describing ballistic trajectories, can get in touch with eyes, nose or mouth of a susceptible person and infect her. The second –*aerosol transmission*– is associated to finer droplets, which can be suspended in the air for hours and easily travel with air currents. The strong dependence of COVID-19 infection risk with people proximity suggests that ballistic transmission is the most relevant contribution to person-to-person contamination [11].

The main protection mechanism of face masks is mostly mechanical in nature: droplets are “filtered out” by stopping their motion when interacting with the mask material [12]. It is then desirable to add further mechanisms to the action of face masks against the transmission of harmful micro-organisms. This “functionalization” has been recently implemented using additives ranging from solidified hand soap [13] to graphene nanomaterials [14].

In this article, we propose a third, natural way of functionalizing face masks materials: the addition of Cuban natural clays. Within lamellar materials, clays and clay minerals stand out due to their wide range of adsorptive properties, and even anti-microbial action [28]. Clay minerals [15-19], and especially Cuban natural clays have shown to be harmless to humans and excellent adsorbents, so they have been studied as drug carriers and de-contaminants [20-24]}. Here, we have functionalized the materials found in the three layers of standard surgical face masks with Cuban natural Bent by different protocols involving aqueous clay suspensions. We test the influence of the treatments in the ability of the materials to stop ballistic droplets using the methodology described in [12], and conclude that this capacity is actually enhanced for the material in the inner layer (i.e., that closer to the user’s face) after the functionalization process. We are currently performing tests to examine the possible enhancement of the microbicidal features of the clay-treated materials.

### Experimental

Three-layer surgical face masks were examined in the assays, due to their widespread use and standard quality (Disposable medical mask model TD-C06, produced by XiantaoTongda Non-Woven Products, Ltd.). Square samples of 5-cm side each were extracted from the masks, corresponding to the inner (INNER), middle (MIDDLE) and outer layers (OUTER).

Each sample was put in contact with aqueous solutions of Ben [20] consisting in 10 g or 20 g of the clay in 400 mL of distiller water, adding either 10 or 20 drops of the cationic surfactant Benzalconium chloride (BC) with magnetic stirring at 500 rpm, during 1 hour, and room temperature. Then, the samples were dried in air also at room temperature for 24 hours. The optical microscopy images were taken with a NOVEL binocular microscope (Digital Light Mobile Phone Novel microscope model XTL-100, produced by Nanbei Instrument Ltd.), to which a Samsung Galaxy A12 cell phone was coupled through an extra convergent lens, acquiring images with a resolution of 3000 × 4000 pixels.

We also quantified the ability of the virgin and functionalized materials to stop ballistic droplets mimicking those produced by human sneezing, for example. To that end, we followed the protocol reported in [12]. Fig. 1 sketches the configuration of the experiment used for the ballistic blocking tests.

**Figure 1.**
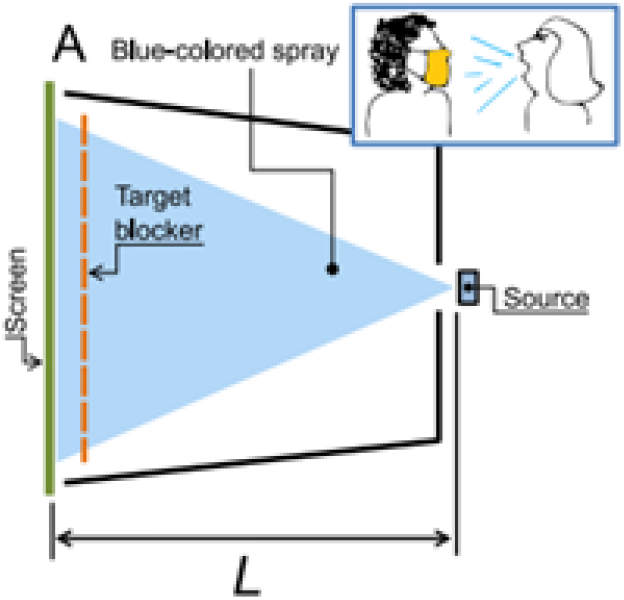
Sketch of the experimental setup used for the calculation of the Ballistic Blocking Capacity (BBC) [12].

The surgical mask samples were fixed in the position “target blocker”, and they were sprayed from a “source” using a methylene blue solution in water (0.5 g/L). A “screen” consisting in a white sheet of paper received the impact of the blue-colored droplets that were not blocked by the target. So, the less stains in the screen, the higher the ballistic blocking capacity of the target under analysis. In order to quantify this effect, we removed the screen after each experiment, which was scanned with a resolution of 3000 dpi. The resulting image was binarized and digitally analyzed in order to calculated the so-called Ballistic Blocking Capacity, defined as:

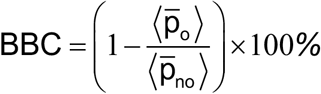

where 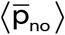 is the average of pixels in the binarized image corresponding to the screen sprayed with no target in place, averaged among all repetitions of the experiment (3, in our case), and 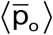 is the analogous parameter, but with a simple in the target position (3 repetitions were also done for each sample). Obviously, since the two parameters can move within the range 0 and 1, BBC = 100% corresponds to a complete stop of all droplets by the simple (completely clean screen, 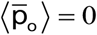), while BBC = 0% indicates that all the droplets managed to pass through the sample, i.e. 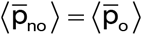).

## Results and discussion

Figure 1 shows the micro-photographs of the functionalized surgical mask samples resulting from the treatments with 10 and 20 g of Bent, and either 10 or 20 drops of surfactant, which will be called “10 DROPS” and “20 DROPS”, respectively. The photos display that the INNER and OUTER layers of the mask are made of lightly-packed polymeric fibers, while the MIDDLE later is made of tightly packed fibers, which is presumably responsible for most of the filtration power of the face mask.

The images suggest that our functionalization protocol results in a homogeneous impregnation of the fibers by the clay, which is revealed by a homogeneous brownish-coloring of the material in the images in rows from 2 to 5 of Fig. 1.

Here we measured the BBC for the INNER sample (i.e., the one that immediately receives the droplets generated by the wearer) before functionalization, and for four different functionalization treatments: F1 (10 g Bent and 10 drops of BC), F2 (10 g Bent and 20 drops of BC), F3 (20 g Bent and 10 drops of BC) and F4 (20 g Bent and 20 BC). The images are shown in Figure 2. Before functionalization, the INNER material showed a BBC of 99.576%. After the different treatments, this material gave the following values: BBC_F1_=99.993%, BBC_F2_=99.998%, BBC_F3_=99.759% and BBC_F4_=99.829%. These results consistently indicate an improvement in the ability to block ballistic droplets after functionalization, especially after the addition of a surfactant.

**Figure 2.**
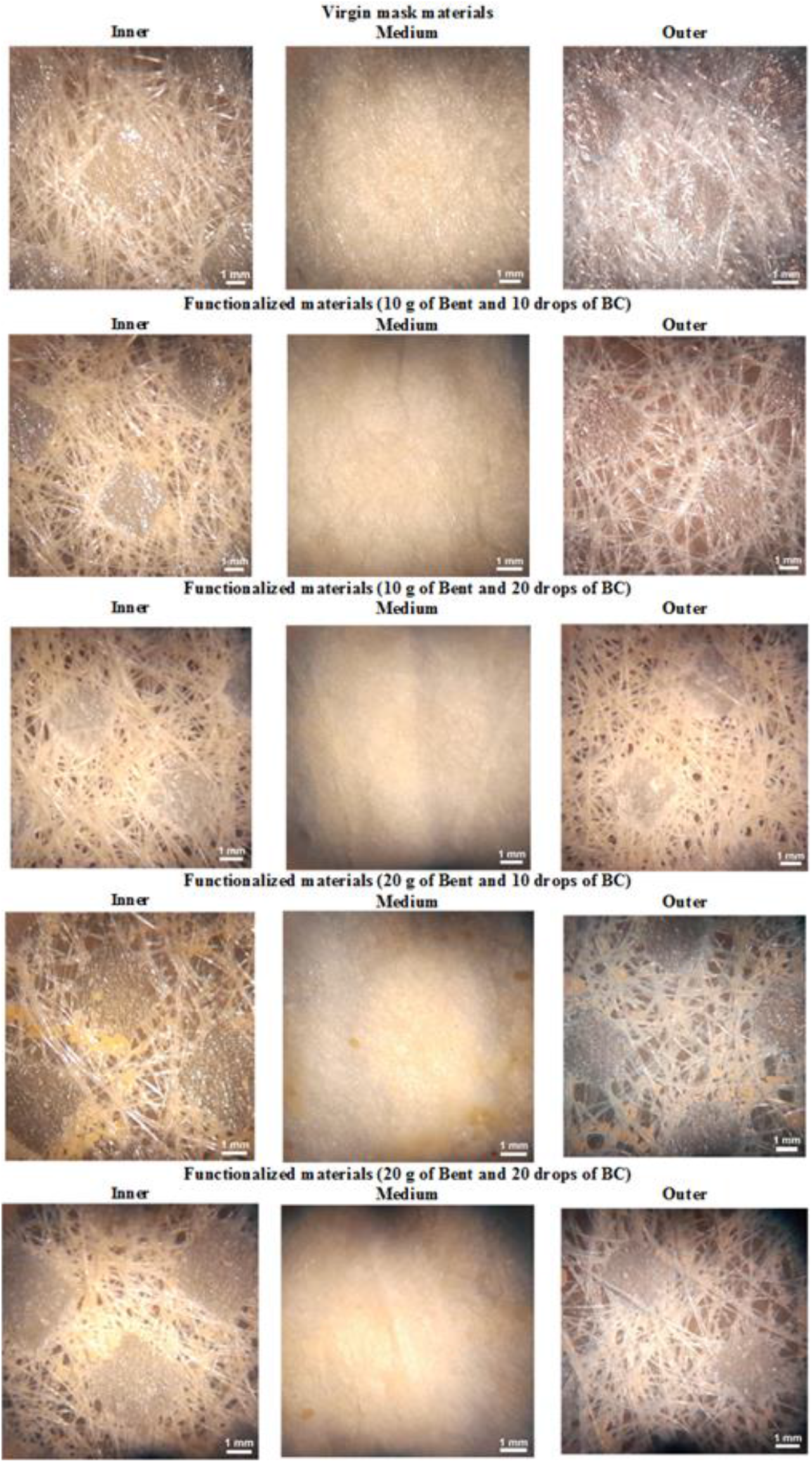
Micro-photographs of surgical mask materials, after and before functionalization. Upper row: Raw samples taken from the surgical masks, with no treatment. Intermediate row: Analogous materials, after functionalization using a clay suspension (10 and 20 g of Bent) in water plus 10 and 20 drops of the benzalkonium chloride (BC) cationic surfactant.

**Figure 3.**
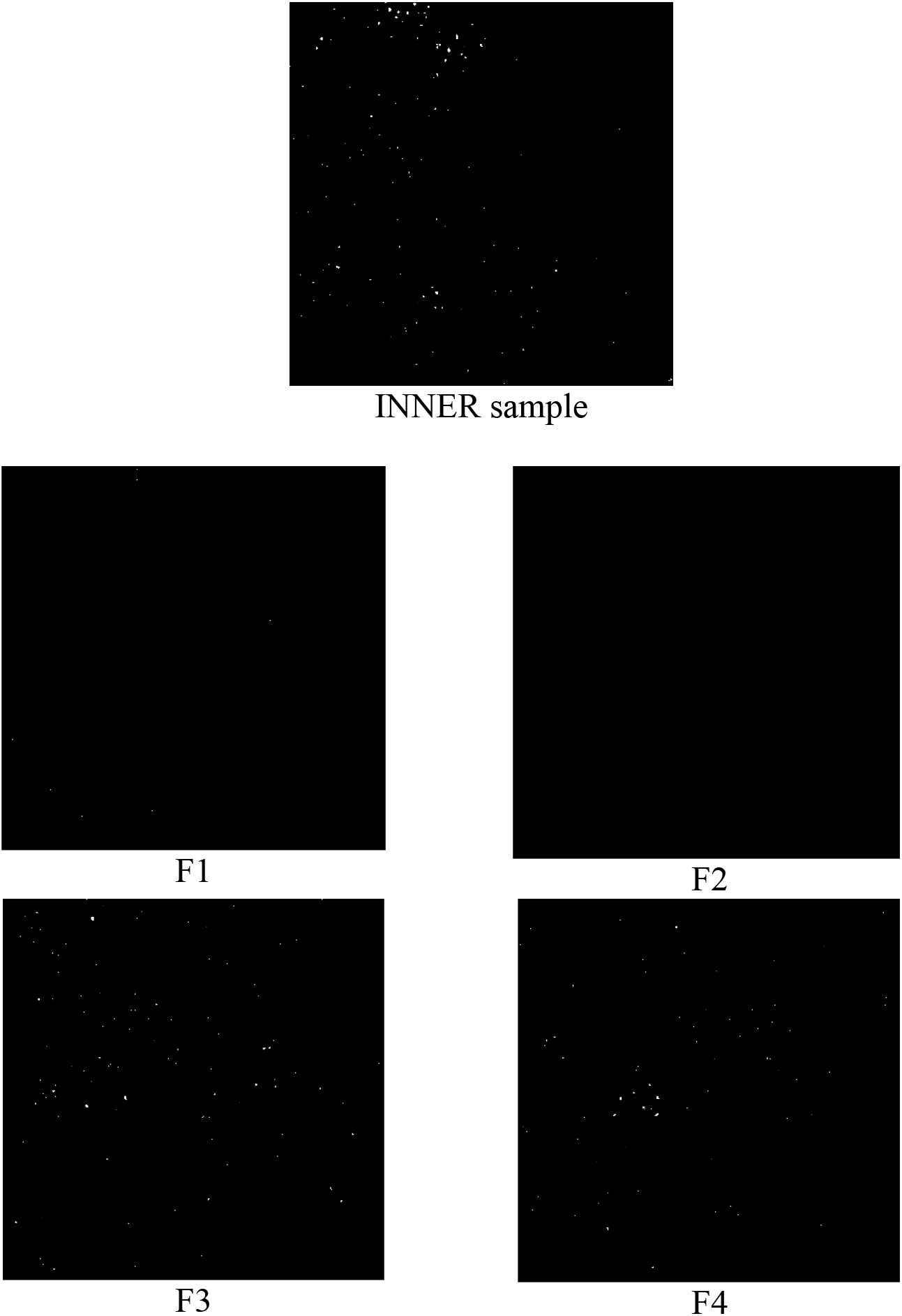
Examples of liquid droplet stains in the screen for the INNER sample (without functionalizing) and four different functionalization treatments (changing the amounts of Bent and CB). These images, that are used to compute the BBCs, have been digitally processed from the original scans.

It is possible to think that this is an expected result since the simple addition of mass to the mask materials should naturally improve their droplet blocking capacity. However, this fact cannot be taken for granted: after all, the strong agitation involved in the functionalization process could have damaged the materials’ fibers.

## Conclusions

Our preliminary results have three main implications:

a. The materials used in standard surgical masks, even without any treatment, effectively block a large percent of the ballistic droplets impacting them (i.e., droplets with sizes larger than 5-10 microns).
b. A simple functionalization treatment with the Cuban natural clay Bentonineis able to incorporate clay particles in all three layers of a standard surgical mask. The inclusion of an ionic surfactant helps that goal.
c. The functionalized materials are not mechanically damaged by the stirring involved in the functionalization process: on the contrary, their ability to block ballistic droplets similar to the ones emitted by human sneezing or coughing is increased after clay incorporation.

It is important to clarify that our study does not shed light on the effect of the functionalization on the stopping of aerosols formed by smaller droplets. However, the fact that the natural clay used has potential anti-microbial properties should weaken the infectious charge in all droplets, including aerosols. The microbiological tests needed to evaluate this possibility are currently being performed.

## Data Availability

All data produced in the present work are contained in the manuscript

## Acknowledgments

V. Marquez is gratefully acknowledged for helping with the BBC determination procedure.

## References

[1] D. K. Chu, E. A. Akl, S. Duda, K. Solo, S. Yaacoub, and H. J. Schünemann, Lancet (London, England) 395, 1973 (2020).

[2] S. Verma, M. R. Dhanak, and J. Frankenfield, Physics of Fluids 32 (2020).

[3] A. Agrawal and R. Bhardwaj, Physics of Fluids 33, 031704 (2021).

[4] W. Lyu and G. Wehby, Health Affairs 39, 10.1377/hlthaff (2020).

[5] J. Howard et al., Proceedings of the National Academy of Sciences of the United States of America 118 (2021).

[6] J. Xi, X. A. Si, and R. Nagarajan, Physics of fluids (Woodbury, N.Y. : 1994) 32, 123312 (2020).

[7] J. Robinson, I. Rios de Anda, F. Moore, J. Reid, R. Sear, and C. Royall, Physics of Fluids 33, 043112 (2021).

[8] D. Maggiolo and S. Sasic, Physics of Fluids 33, 083305 (2021).

[9] A. Konda, A. Prakash, G. A. Moss, M. Schmoldt, G. D. Grant, and S. Guha, ACS nano 14, 6339 (2020).

[10] T. Dbouk and D. Drikakis, Physics of fluids (Woodbury, N.Y. : 1994) 32, 063303 (2020).

[11] E. A. Meyerowitz, A. Richterman, R. T. Gandhi, and P. E. Sax, Annals of internal medicine 174, 69 (2021).

[12] V. Márquez-Alvarez, J. Amigó-Vega, A. Rivera, A. J. Batista-Leyva, and E. Altshuler, PLOS ONE 17, e0275376 (2022).

[13] A. Cano-Vicent, A. Tuñón-Molina, M. Martí, Y. Muramoto, T. Noda, K. Takayama, and A. Serrano-Aroca, ACS Omega 6, 23495 (2021).

[14] F. De Maio et al., iScience 24, 102788 (2021).

[15] A. Rivera et al., Appl. Clay Sci. 124, 150 (2016).

[16] D. Hernández, L. Lazo, L. Valdes, L. C. de Ménorval, Z. Rozynek, and A. Rivera, Appl. Clay Sci. 161, 395 (2018).

[17] L. Valdés, D. Hernández, L. C. de Ménorval, I. Pérez, E. Altshuler, J. O. Fossum, and A. Rivera, Eur. Phys. J. Spec. Top. 225, 767 (2016).

[18] L. Valdés, S. A. Martín, D. Hernández, L. Lazo, L. C. de Ménorval, and A. Rivera, Rev. Cub. Fís. 34, 35 (2017).

[19] L. Valdés, I. Pérez, L. C. de Ménorval, E. Altshuler, J. O. Fossum, and A. Rivera, PlosOne 12, e0187879 (2017).

[20] D. Hernández et al., Pharmaceutics 15, 1171 (2023).

[21] D. Hernández, L. Quiñones, L. Lazo, C. Charnay, M. Velázquez, E. Altshuler, and A. Rivera, Journal of Porous Materials 30, 1149 (2022).

[22] S. A. Martín, I. Pérez, and A. Rivera, Applied Clay Science 202, 105965 (2021).

[23] D. Hernández, L. Quiñones, C. Charnay, M. Velázquez, and A. Rivera, Rev. Cubana Fis. 36, 144 (2019).

[24] S. A. Martín, L. Valdés, F. Mérida, L. C. de Ménorval, M. Velázquez, and A. Rivera, Clay Miner. 53, 193 (2018).

